# Lower RNA expression of ALDH1A1 distinguishes the favorable risk group in acute myeloid leukemia

**DOI:** 10.1101/2021.10.20.21265241

**Authors:** Garrett M. Dancik, Ioannis F. Voutsas, Spiros Vlahopoulos

**Affiliations:** Department of Computer Science, Eastern Connecticut State University, Willimantic, CT USA.; Cancer Immunology & Immunotherapy Center, Saint Savas Cancer Hospital, 171 Alexandras Avenue, 11522, Athens, Greece; First Department of Pediatrics, National and Kapodistrian University of Athens, Thivon & Levadeias 8,11527 Goudi-Athens, Greece

**Keywords:** Aldehyde Dehydrogenase, Drug Resistance, Immunosuppression, Leukemia, Myeloid, Acute, Neoplastic Stem Cells, Gene Expression, Biomarkers

## Abstract

The expression and activity of enzymes that belong to the aldehyde dehydrogenases is a characteristic of both normal and malignant stem cells. ALDH1A1 is an enzyme critical in cancer stem cells. In acute myeloid leukemia (AML), ALDH1A1 protects leukemia-initiating cells from a number of antineoplastic agents, which include inhibitors of protein tyrosine kinases. Furthermore, ALDH1A1 proves vital for the establishment of human AML xenografts in mice. We review here important studies characterizing the role of ALDH1A1 in AML and its potential as a therapeutic target. We also analyze datasets from leading studies, and show that decreased ALDH1A1 RNA expression consistently characterizes the AML patient risk group with a favorable prognosis, while there is a consistent association of high ALDH1A1 RNA expression with high risk and poor overall survival. Our review and analysis reinforces the notion to employ both novel as well as existing inhibitors of the ALDH1A1 protein against AML.

## 1. Introduction

Acute myeloid leukemia is a clonal hematopoetic disease that is associated with a high death rate in pediatric (>30%) as well as in adult (>50%) patients. In adults AML is the most common form of acute leukemia, and has the shortest survival, with less than 25% of patients surviving five years after diagnosis [1]; [2]; [3]. AML can be classified into primary, de novo AML, which arises in the absence of an identified exposure or prodromal stem cell disorder and non-denovo AML; non-denovo includes secondary AML, representing transformation of an antecedent diagnosis of myelodysplastic syndrome (MDS) or myeloproliferative neoplasm, therapy-related AML developing as a late complication in patients with prior exposure to leukemogenic therapies [4]. From a biological and therapeutic standpoint, relapsed AML should also be included in non-de novo AML, as it involves discrete, traceable changes [5] from primary AML.

Although AML is genetically heterogeneous, it involves a rather small number of genetic alterations and certain molecular patterns prove common to both adult as well as to pediatric patients [6]; [7]. The World Health Organization defines specific acute myeloid leukemia (AML) disease entities by focusing on significant cytogenetic and molecular genetic subgroups: a large number of recurring, balanced cytogenetic abnormalities are recognized in AML, and most of those that are not formally recognized by the classification are rare [8].

### 1.1. Classification of AML into prognostic risk groups

The primary prognostic factor in AML is cytogenetics, due to the fact that the clinical course of AML is largely determined by the presence of specific chromosomal aberrations in the cell nucleus. Depending on the types of chromosomal aberrations present, AML patient cases were classified into low risk, intermediate risk, and high risk. Cases with no detected chromosomal aberrations are classified into the intermediate risk group, while cases with detected aberrations are classified into any one of the three categories. Specifically AML associated with t(8;21), t(15;17) or inv(16) predicted a relatively favorable outcome. Whereas in patients lacking these favorable changes, the presence of a complex karyotype, -5, del(5q), -7, or abnormalities of 3q defined a group with relatively poor prognosis. The remaining group of patients including those with 11q23 abnormalities, +8, +21, +22, del(9q), del(7q) or other miscellaneous structural or numerical defects not encompassed by the favorable or adverse risk groups were found to have an intermediate prognosis [9].

In 2010, an international expert panel, on behalf of the European LeukemiaNet (ELN), published recommendations for diagnosis and management of acute myeloid leukemia (AML), which integrated cytogenetics with mutational screening for the genes NPM1, FLT3, and CEBPA that improved classification. The risk groups were defined as low-risk, intermediate I, intermediate II, and high-risk (or with reference to prognosis as favorable, intermediate 1, intermediate 2, and adverse) [10].

These recommendations have been widely adopted in general practice, within clinical trials, and by regulatory agencies. The original intention of the ELN genetic categories was to standardize reporting of genetic abnormalities particularly for correlations with clinical characteristics and outcome.

During recent years, considerable progress has been made in understanding disease pathogenesis, and in development of diagnostic assays and novel therapies. In 2017, the panel decided to simplify the ELN system by using a 3-group classification (favorable, intermediate, adverse) rather than the previous 4-group system; in parallel, mutational screening for genes RUNX1, ASXL1, and TP53 was added to the prognostic criteria [11].

### 1.2. Cellular developmental changes affect the response to treatment

Even though AML was among the first malignancies to show durable remission with adoptive therapy though transplantation of healthy stem cells, there is a paucity of information on the effects of therapeutic agents on key factors that shape the leukemia microenvironment [12]. Furthermore, there is a need of a deeper molecular understanding that would lead to treatments targeted to the leukemia-initiating cells (or “leukemia stem cells”) that appear linked to the most severe and difficult to treat manifestations of AML [13].

The myeloid progenitor cells arise from hematopoietic stem cell by asymmetric division. In turn the myeloid progenitor cells give rise to myeloblasts. An abnormal clone that develops from either a myeloid progenitor cell or a myeloblast can give rise to acute myeloid leukemia [14]. Characteristically, internal tandem duplications of the FLT3 gene are present in AML stem cells, and support key leukemia-initiating cell properties, which are experimentally shown by engraftment into mice [15]. Patients with AML cells expressing a primitive stem cell phenotype (CD34+CD38-with high aldehyde dehydrogenase activity) manifest significantly lower complete remission rates, as well as poorer event-free and overall survivals [16]. In AML, signaling from the bone marrow microenvironment of normal hematopoietic stem cells and leukemia-initiating cells has a pivotal role in the biology of the disease [17]. Deficient signaling from the vitamin A derivative, all-trans retinoic acid in the hematopoietic microenvironment in mice was shown to cause a myeloproliferative syndrome with significant reduction in the numbers of B lymphocyte subsets and erythrocytes in the bone marrow [18].

In patients with acute promyelocytic leukemia, high blast cell counts and failure to respond to differentiation treatment was associated with low all-trans retinoic acid plasma concentrations, and a high rate of clearance [19]. For the other types of AML, and despite its importance in controlling myeloid differentiation and apoptotic pathways, all-trans retinoic acid has yet to prove itself as a useful agent in the armamentarium of AML therapeutics. The explanation probably lies in the fact that retinoic acid receptor is expressed, but of limited functionality in AML blast cells [20]. However there are both positive and negative associations of CD34+ immature blast cells with sensitivity to all-trans retinoic acid [21]; [22].

## 2. Aldehyde dehydrogenases in AML

### 2.1. Implication of the enzymatic activity of aldehyde dehydrogenases in AML development and progression

Aldehyde dehydrogenases are enzymes that oxidize aldehydes to carboxylic acids. Member proteins of the subfamily ALDH1 metabolize the retinaldehyde, the product of oxidation of vitamin A, into retinoic acid, which binds to the retinoic acid receptors α, β, and γ, to activate them as regulators of gene expression. This function helps differentiation of diverse cell types, and primes tissues for repair, regrowth and regeneration after inflammation or injury, and activates genes of the HOX family, which in turn activate cellular programs of positional memory during embryogenesis and regeneration [23].

In the organism, ethanol is metabolized to acetaldehyde that interferes with systems that protect cells from oxidant stress. ALDH proteins oxidize acetaldehyde to acetate, protecting cells and tissues from oxidative damage and helping maintain tissue function. However, ALDH proteins protect both normal as well as malignant stem cells (cancer stem cells) from reactive aldehydes and certain cytotoxic drugs such as cyclophosphamide [24]; [25].

Evidence has been mounting that the protein family of the aldehyde dehydrogenases plays a role in the development of AML, although the interpretation of their exact biological effect on AML progression remains controversial [26]. Higher ALDH activity was shown in normal hematopoietic stem cells, when compared to AML stem cells in a study of 32 patients and five bone marrow donors [27]. In a larger sample size (n=104), a minority of patients (24 of 104) had numerous ALDH-positive leukemic stem cells that could not be separated from normal hematopoietic stem cells, were drug-resistant, and had high efficiency of xenograft formation in mice [28]. Nevertheless, cases with increased fractions of ALDH-positive AML cells were shown to derive from immature hematopoietic progenitors, suggesting an explanation for the poor prognosis and therapy resistance of this subgroup, which was attributed to the transmission of stem cell properties [29][30].

### 2.2. Overview and function of ALDH1A1

ALDH family member ALDH1A1, encoded in chromosome 9q21, is the enzyme of the second step of the main oxidative pathway of alcohol metabolism; ALDH1A1 may also catalyze the oxidation of toxic lipid aldehydes such as 4-hydroxynonenal and malonaldehyde, and is the enzyme responsible for production of retinoid acid during the late stages of dorsal eye development [31]. Notably, citral and disulfiram, an FDA-approved drug to treat alcohol use disorder, inhibit human lens ALDH1A1 at IC50 values of the micromolar range [32]. In neoplasia, in addition to its known enzymatic activity, ALDH1A1 supports tumor growth via glutathione/dihydrolipoic acid-dependent NAD + reduction [33]. ALDH1A1 is elevated in hematopoietic stem cells, is a predominant ALDH isoform in mammalian tissues, and the key ALDH isozyme linked to both normal and malignant stem cells [34]. ALDH1A1 in hematopoietic stem cells metabolizes reactive aldehydes and reactive oxygen species, and has a potential role in protection of neurons from 3,4-dihydroxyphenylacetaldehyde [35]. Consequently, decreased expression of ALDH1A1 has been linked to the development of Parkinsons’ disease [36].

Furthermore, the ALDH1A1 protein is a critical enzyme for maintaining clarity in human, rat, and mouse lenses; ALDH1A1-null mice grow to adulthood, but develop cataracts later in life (by six to nine months of age;) [37]; [38]; [32]. Hematopoietic cells from ALDH1A1-deficient mice exhibit increased sensitivity to liver metabolites of cyclophosphamide; however, ALDH1A1 deficiency did not affect the function of stem cells from the hematopoietic and nervous system [39].

### 2.3. Regulation of ALDH1A1 gene expression

Expression of the gene ALDH1A1 is regulated by the transcription factor EVI1 (Ecotropic virus integration site-1), which has an outstanding role in the formation and transformation of hematopoietic cells; a meta-analysis suggested that EVI1 is associated with an adverse prognosis in AML, and specifically in patients that belong to the intermediate risk group according to cytogenetic criteria of the European LeukemiaNet (ELN) [40]; [41]; [42]. Another transcription factor implicated in ALDH1 regulation is nuclear factor kappa B (NFκB) a factor found constitutively active in malignant myeloblast cells of AML [43]; [44]; [45]. Cells normally activate NFκB in response to inflammatory stimuli and disruptions of tissue function [46].

To activate ALDH1A1 expression indirectly, NFκB induces expression of micro RNA223-3p, which inhibits expression of ARID1A. ARID1A loss, in turn allows histone acetylation of the ALDH1A1 gene promoter [47]; [48]. Having an established capacity to guide epigenetic changes that guide leukemic stem cell programs, NFκB has been associated with cancer recurrence, and with AML relapse [49]; [50]; [51]; [52].

### 2.4. An elusive role for ALDH1A1 expression in AML

In AML the expression of RNA from the ALDH1A1 gene was implicated in resistance of AML to the protein kinase inhibitor sorafenib in FLT3-mutated AML, which normally shows potent FLT3-ITD (internal tandem duplication) inhibitory activity [53] ; [54]. Specifically, nonobese diabetic/severe combined immunodeficiency mice (NOD/SCID) transplanted with leukemia cells from patients before and during sorafenib resistance recapitulated the clinical results: The CD34+ALDH+ fraction preferentially expressed ALDH1A1 and exhibited superior engraftment compared with the CD34+ALDH-population when transplanted to NOD/SCID mice at equal cell doses. Forced expression of ALDH1A1 enhances the colony-forming capacity of leukemia cell lines 697(EU3) and HL60/S4 [55].

ALDH1A1 overexpression has also been associated with sorafenib resistance in other malignancies [56]. Interestingly however, in a study of primary AML cells from 21 patients, ALDH1A1 transcripts were highest in the ALDHneg leukemic cells [57]. On the other hand, in mouse xenografts, use of an irreversible inhibitor of ALDH1 eradicated specifically human AML cells and spared healthy mouse hematologic cells; in vitro, selectively the inhibitor was cytotoxic in a leukemic population enriched in stem cells, but unlike conventional chemotherapy, it was not toxic for healthy hematopoietic stem cells [58]. Furthermore, increased expression of ALDH1A1 was shown to confer to AML cells resistance to reactive aldehydes and reactive oxygen species, and associated with a worse prognosis in AML patients of The Cancer Genome Atlas (TCGA) study [59].

From all of the above data it can be concluded that the ALDH gene family has been implicated in the biology of AML; however, data on the important isoform ALDH1A1 have not been entirely conclusive in regard to the exact effect of ALDH1A1 expression on AML prognosis. We therefore sought to characterize the relationship between ALDH1A1 expression, cytogenetic risk group, and prognosis by examining publicly available datasets.

## 3. Cross-dataset analysis

### 3.1. Patient cohorts and data processing

We searched the literature and the Gene Expression Omnibus [60] for publicly available AML gene expression datasets that included risk or survival information. We identified 8 cohorts with risk information (N = 1473) and 7 with overall survival information (N = 1170). The cohorts we analyzed are summarized in **Table 1**. Two datasets, GSE12417 and GSE10358, were identified but were excluded from our analysis because they contain duplicate data. In particular, GSE12417 contains patients from the AMLCG trial which are profiled in GSE37642, and GSE10358 contains data from TCGA. We note that we limited our analysis to differential expression (across risk groups) and overall survival analysis since other variables (such as treatment assignment and response) were not consistently reported.

**Table 1.** Summary of patient cohorts

| Cohort | Description | Reference (Pubmed ID) |
| --- | --- | --- |
| TCGA | Adult patients with de novo AML with expression profiles available through The Cancer Genomic Atlas (TCGA) | 23634996 |
| TARGET | Samples from pediatric patients from the NCI/COG TARGET-AML initiative. | 26941285 |
| BEAT AML (BMA) | Bone marrow aspirate (BMA) or peripheral blood (PB) primary tumor samples from the BEAT AML program. Patients with both BMA and PB samples are excluded. | 30333627 |
| BEAT AML (PB) |  |  |
| GSE37642 (GPL570) | Two independent patient cohorts from the German AMLCG 1999 trial, profiled on the Affymetrix Human Genome U133A array (GPL96) or the U133 Plus 2.0 Array (GPL570) | 23382473 |
| GSE37642 (GPL96) |  |  |
| GSE6891 (Cohort #1) | De novo AML samples from two independent cohorts. Cohort #1 includes samples below 4000; Cohort #2 includes samples above 4000. Samples added in 2011 are excluded. | 18838472 |
| GSE6891 (Cohort #2) |  |  |
| GSE71014 | De novo AML samples from cytogenetically normal patients | 26517675 |

RNA-seq count data was downloaded for the TCGA and TARGET cohorts. TCGA data was downloaded from Firehose [61]. TARGET expression data was downloaded from the Genomic Data Commons [62]) and clinical information downloaded from cBioPortal [63]. RNA-seq count data was processed using *R*. First, low expressing genes (<15 read counts across samples) were removed. Counts were normalized using the TMM method using the *edgeR* package [64], and then converted to counts per million (CPM) on the log scale [65]. BEAT AML data was imported into *R* using the *beatAML* package (https://github.com/radivot/AMLbeatR) and filtered to include only initial leukemia samples. The dataset was then split into two cohorts, for patients with bone marrow aspirate (BMA) samples and patients with peripheral blood (PB) samples. Patients with both BMA and PB samples (N = 17) were excluded from downstream analysis, so that the BMA and PB cohorts could be analyzed independently. All other expression data was downloaded from the Gene Expression Omnibus [60] and analyzed using *R*.

### 3.2. AML patient risk groups are associated with overall survival

We first confirm that assigned risk groups are associated with patient outcomes by generating Kaplan-Meier curves for the 6 cohorts with both survival and risk information (**Figure 1**). Risk status was determined using ELN 2017 criteria with labels provided by the original authors. In 5/6 cohorts, risk status is significantly able to stratify patients (logrank P < 0.01), with the patients in the “favorable” or “low” risk group having a longer survival. In one cohort, BEAT AML (PB), the result is not statistically significant, but patients in the “favorable” risk group generally have better outcomes. These results demonstrate that, as expected, risk status is associated with outcome in the cohorts we examine.

**Figure 1.**
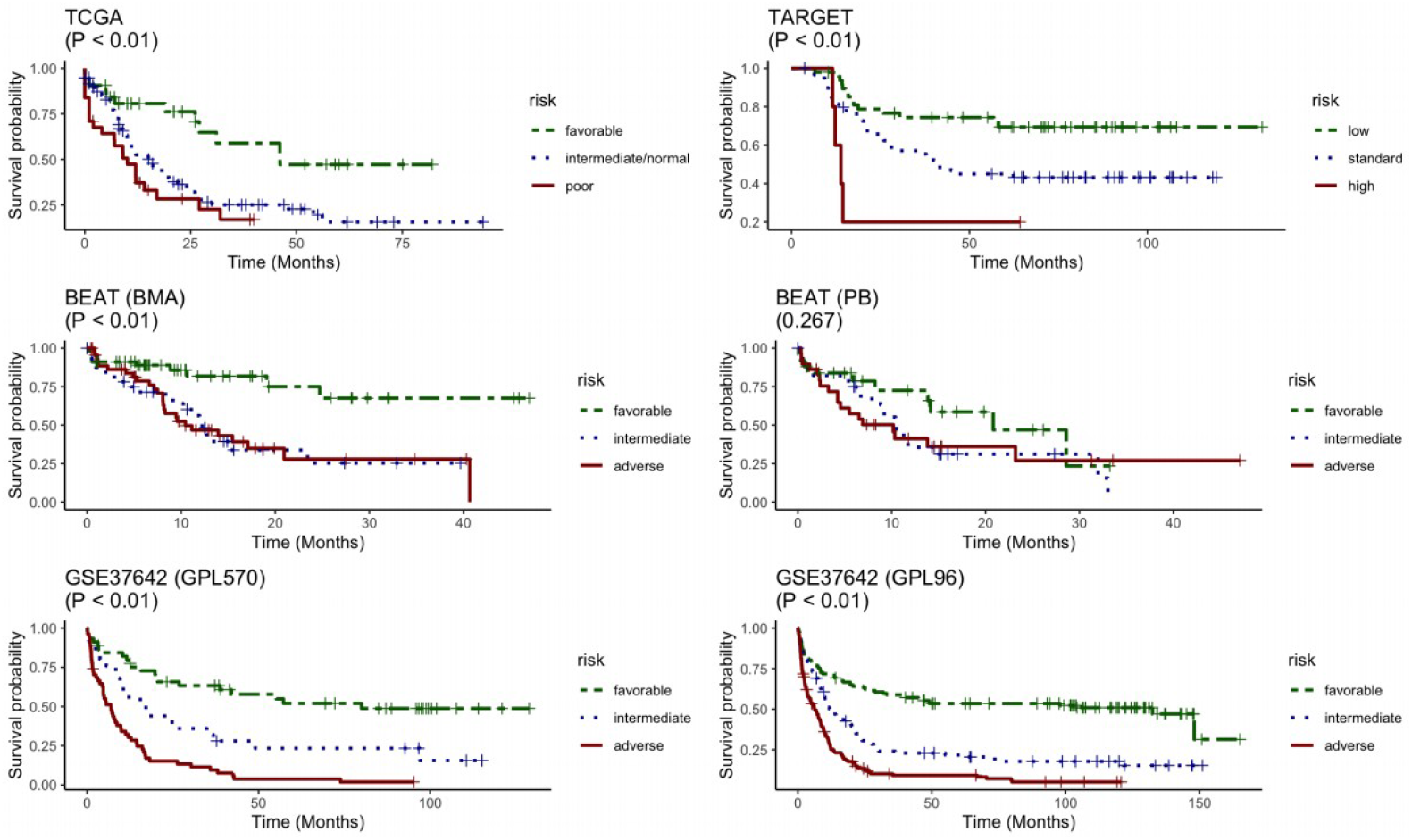
Kaplan-Meier curves of overall survival stratified by patient risk groups across patient cohorts. Cohorts include TCGA (N = 159), TARGET (N = 113), BEAT (BMA) (N = 135), BEAT (PB) (N = 83), GSE37642 (GPL570) (N = 124), and GSE37642 (GPL96) (N = 370). P-values are calculated using the log-rank test.

### 3.3. ALDH1A1 RNA expression levels in AML patient risk groups

We next examined the relationship between ALDH1A1 RNA expression level and risk group, and find that expression consistently differs across risk groups (P < 0.01) in 8 patient cohorts. In all cases, ALDH1A1 expression is the lowest in the “favorable” or “low” risk group (**Figure 2**).

**Figure 2.**
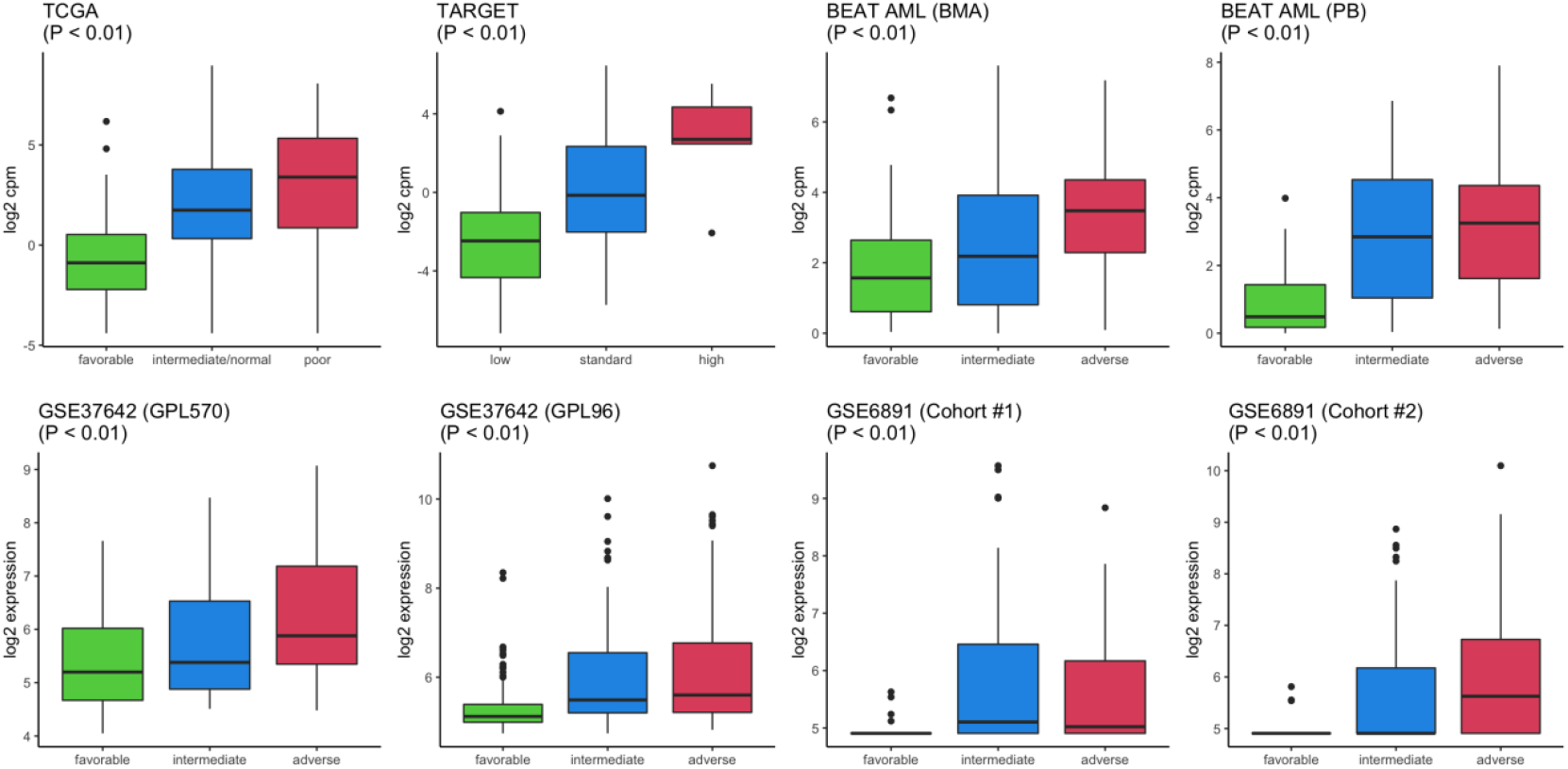
Association of ALDH1A1 expression and risk groups across patient cohorts. Cohorts include TCGA (N = 171), TARGET (N = 113), BEAT (BMA) (N = 150), BEAT (PB) (N = 93), GSE37642 (GPL570) (N = 124), GSE37642 (GPL96) (N = 371), GSE6891 (Cohort #1) (N = 244), and GSE6891 (Cohort #2) (N = 207). P-values calculated by ANOVA.

### 3.4. ALDH1A1 RNA association with AML patient overall survival

A survival analysis was carried out to determine whether ALDH1A1 expression was associated with overall survival of AML patients. For each cohort, patients were stratified based on whether their ALDH1A1 expression level was high (**≥** median) or low (< median). A forest plot summarizing the survival analysis for the 7 cohorts is given in **Figure 3**. In each case the HR > 1, indicating that low expression is consistently associated with a more favorable outcome, and this relationship is statistically significant in 3 cohorts (P < 0.05). Additionally, the HR has a weighted average of 1.42 (95% CI, 1.22 – 1.66), which is statistically significant (P < 0.05).

**Figure 3.**
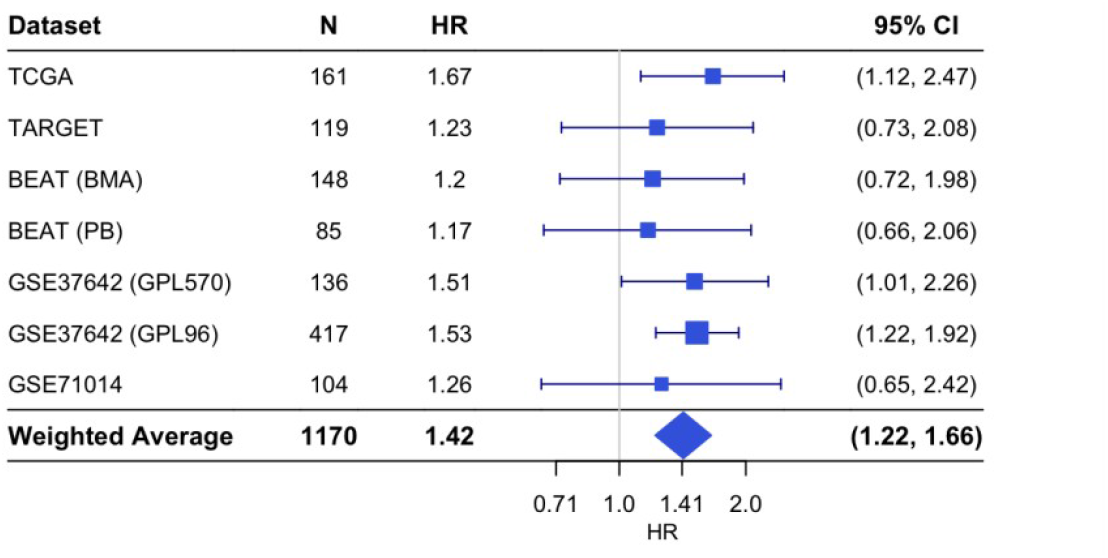
Forest plot of hazard ratios for overall survival in patient cohorts based on ALDH1A1 expression. Each hazard ratio (HR) is calculated by comparing survival curves for patients with high ALDH1A1 expression (**≥** median) to patients with low (< median) expression. HR > 1 corresponds to patients with high expression having a higher risk. For each individual cohort, the HR and 95% confidence interval (CI) is denoted by the blue rectangle and whiskers, respectively. The size of the blue rectangle is proportional to the precision of the HR estimate. For the weighted average, the diamond represents the 95% CI.

## 4. Discussion

### 4.1. Biological mechanisms associated with ALDH1A1 suggest a path for new treatment strategies

Expression of the ALDH1A1 gene, and the consequent activity of the resulting ALDH1A1 protein influence a number of distinct biological functions, either directly or indirectly. ALDH1A1 is an essential enzyme for development and maintenance of tissue homeostasis, yet its overexpression is evidently linked to the development and progression of AML. We here discuss a few representative phenomena that implicate ALDH1A1 gene expression, with focus on functions that have a marked impact on AML disease course.

### 4.2. ALDH1A1 activation of the myeloid stem cell lineage associates with severe disease course

A critical discovery was made when it was demonstrated that overexpression of ALDH1A1 promoted myelopoiesis, while inhibiting lymphopoiesis in murine hematopoietic progenitors. This finding can explain the uniform association of ALDH1A1 overexpression with the poor prognosis in AML, as it could be interpreted with the hypothesis that in poor-prognosis AML, ALDH1A1 RNA is expressed far above the level that is required for the myeloid lineage. This overexpression could be due to higher activity of transcription factors such as TLX1/HOX11 and alternatively, NFκB [66]. Dysregulation of either one of these factors has been associated with a severe course of AML [67]; [68]. In particular NFκB has been characterized as a factor with an important role in the control of AML stem and progenitor cells, with a vital function in the regulation of interactions between AML cells and their microenvironment [69]; [70]; [52]; [71].

### 4.3. AML stem cells are maintained by signaling through FLT3 and NFκB

In AML, constitutive NFκB DNA-binding activity is frequently mediated by a Ras/PI3-K/PKB-dependent pathway [72]. Also overexpression of FLT3 induces NFκB-dependent transcriptional activity in cultured cells [73]. The constitutive activation of FLT3-ITD induces NFκB activity [74]. And conversely, FLT3 inhibition or knockdown reduces constitutive NFκB activation in high-risk myelodysplastic syndrome and AML [75]. NFκB is regulated by changes in cellular proteostasis; its native inhibitor, the IkB protein can be degraded by cellular proteolytic systems such as the proteasome or the lysosome, which are mutually regulated through the induction of transcription factors [76]; [77]. In fact, induction of cell stress combined with NFκB inhibition is a promising approach in AML treatment, due to the connection between proteostatic pathways activated by cell stress and those inducing NFκB activity, as was recently shown in a clinical trial of relapsed patients with acute promyelocytic leukemia [78]; [79].

One agent that is used in treatment of AML, is the nucleoside cytosine arabinoside [80]. In fact, cytosine arabinoside (cytarabine, ara-C), in high concentrations (>50uM) inhibits NFκB in some cell types to a certain extent; yet this inhibition is evidently not enough, since after development of drug resistance, ara-C does not suffice as monotherapy for AML [81]; [82]; [83].

On the one hand, mutated, constitutively active FLT3, activates signaling molecules RAS, and PI3K/mTOR which activate NFκB [84]. On the other hand, AML can develop resistance against all inhibitors of FLT3 at some point [85]. Consequently, combination of NFκB and FLT3 inhibitors has been previously suggested by in vitro and xenograft studies of AML cells from patients [86]; [87]. An additional reason to combine inhibitors is the fact that NFκB causes epigenetic changes in nuclear chromatin, which cannot be reversed by inhibition of NFκB itself in the affected cells [88]. Downstream target genes of NFκB-guided transcription, therefore, such as ALDH1A1, are attractive aims for AML treatment.

### 4.4. ALDH1A1 protects malignant cells and modulates effects of AML chemotherapy

ALDH1A1 causes resistance to several antineoplastic treatments. A number of agents, such as paclitaxel, doxorubicin, sorafenib and staurosporine, promote oxidant-stress and thereby cause cell death; both phenomena are inhibited by ALDH1A1 expression [56]. Conversely, under conditions that ALDH1A1 is inhibited, resistance of cancer cells to cytarabine is decreased and cells become sensitive to the drug. For AML, disulfiram was found to suppress cytarabine resistance of cells with high aldehyde dehydrogenase activity, and to inhibit their engraftment in mice, while sparing healthy blood cells [89]. Thus AML is dependent on ALDH1 activity, and more sensitive to ALDH1 inhibition than the normal hematopoietic system.

ALDH1A1 leading to the myeloid lineage would be consistent with an increased lysosomal capacity, and a chemoresistant profile of the resulting cells [90]; [91]; [92]; [93]. In fact, autophagy supplies AML cells with lipid degradation products, which fuel oxidative phosphorylation in a manner that is absent in normal cells [94]. Even inhibition of NFκB conveys ara-C sensitivity more effectively to bulk AML cells than to leukemia stem cells, while stem and progenitor AML gains in resistance, presumably via lysosomal pathway activation: this is shown by the synergy between mTOR inhibitors and autophagy inhibitors in killing AML cells [95]; [96]. However, ALDH1A1 overexpression was found to enhance lysosomal autophagy inhibitor cell entry and cytotoxicity, without directly affecting lysosome function or autophagic flux [97]. Furthermore, inhibition of autophagy did not convert AML cells from ara-c resistance to sensitivity [98]. This can be explained by an increased capacity of ara-c resistant AML to respond to the oxidative DNA damage that is caused by ara-c, and by a contribution of ALDH1A1 to the detoxification of reactive aldehydes that would otherwise saturate the DNA with oxidative lesions.

In fact, relapse in AML was previously associated with increases in advanced oxidation protein products, malondialdehyde, and 8-hydroxydeoxyguanosine, and decreased total antioxidant capacity in patients’ plasma [99]. Constitutively active FLT3, in particular increases reactive oxygen species production leading to increased DNA double-strand breaks and DNA repair errors that may explain aggressive AML [100]. In particular, genomic instability is a principle pathologic feature of AML with internal tandem duplications of the FLT3 gene, which is characterized by poor prognosis [101]. AML cells adapt to oxidant stress by adapting sphingolipid metabolism, fatty acid oxidation, purine metabolism, amino acid homeostasis and glycolysis [102]. However, oxidant stress alters the cellular microenvironment, by inducing the expression of cytokines and chemokines:

Reactive aldehydes and reactive oxygen species generate oxidative damage on DNA, which activates repair enzyme binding to DNA. When increased on the cell, oxidant stress temporarily inactivates base excision repair enzyme 8-oxoguanine glycosylase (OGG1), which recruits transcription factor NFκB to induce expression of inflammatory genes [103]. This effect has profound effects in RAS/NFκB-dependent carcinogenesis, by modulating interactions between cancer cells, their microenvironment, and the immune system, causing immunosuppression [2]; [104]. In hepatocarcinogenesis, fluctuations of ALDH1A1 and NFκB are observed, concomitantly with OGG1 [105]. In AML, OGG1 expression correlates with adverse cytogenetics, and has a more adverse impact on disease outcome in the context of the FLT3-ITD mutation [106], a context shared with ALDH1A1 [54]. In fact, OGG1 affects resistance of AML to cytarabine: OGG1-deficient acute myeloid leukemia cells have increased sensitivity to cytarabine [107].

It can be therefore concluded that reactive oxygen species are essential to the effects of certain chemotherapeutic agents, and increases in ALDH1A1 cause resistance to these agents and facilitate relapse of AML. However, in other cancers ALDH1A1 expression was linked to the expression of PD-L1, a molecule that suppresses T-lymphocyte mediated immunity [108]; [109]. PDL-1 is expressed in the microenvironment of AML, where it promotes expansion of the immunosuppressive regulatory T-cells [110]; [111]; [112]. If the same PD-L1 inducing effect of ALDH1A1 occurs also in AML as in other cancers (**Supplementary Figure S1**), then high ALDH1A1 expression in cases with adverse prognosis may be linked to immune evasion, in addition to the well-characterized effects of ALDH1A1 on antineoplastic agents.

### 4.5. Implications for treatment

Novel selective inhibitors for ALDH1A1 are under development [113]; [114]; [115]; [116]. In other malignancies, clinical trials evaluating the ALDH1A1 inhibitor disulfiram in glioblastoma have been registered (U.S. National Library of Medicine resource platform ClinicalTrials.gov registration numbers NCT01777919, NCT02770378, NCT01907165, NCT03363659, NCT02678975, NCT03151772, NCT03034135, NCT02715609; for an overview see https://clinicaltrials.gov/ct2/results?term=disulfiram&cond=Cancer). Other, experimental inhibitors of ALDH1A1 should enter translational research for the development of AML treatments. A success in this endeavor would be especially important for patients with poor-prognosis, including those experiencing disease relapse and resistance to chemotherapy and biologically targeted therapy.

Additionally, the consistent up-regulation of ALDH1A1 expression in patients with non-favorable risk across a wide range of patient cohorts (**Figure 2**) suggests that such a treatment may benefit a wide variety of patient groups.

## Data Availability

All data produced in the present work are contained in the manuscript.

## Abbreviations

AKT/PKB: RAC-alpha serine/threonine-protein kinase
ALDH: Aldehyde dehydrogenase
AML: Acute myeloid leukemia
ara-c: cytosine arabinoside
CEBPA: CCAAT enhancer binding protein alpha
del: deletion
ELN: European LeukemiaNet
EVI1: ecotropic viral integration site 1
FLT3: Fms-like tyrosine kinase-3
FLT3 ITD: internal tandem duplication of FLT3
inv: inversion
MTOR, kinase: mechanistic/mammalian target of rapamycin
NFκB: nuclear factor kappa B
NOD/SCID: nonobese diabetic/severe combined immunodeficiency mice
NPM1: nucleophosmin 1
OGG1: 8-oxoguanine glycosylase
PD-L1: Programmed death-ligand 1
PI3K: phosphatidylinositol 3-kinase
t: translocation
TLX1/HOX11: T cell leukemia/homeobox 1

## Acknowledgements

Authors wish to thank the AMLCG study group for help in this project. This work was supported, in part, by a grant from the American Association of University Professors and Connecticut State University Board of Regents (YRDA01).

## References

1. Pollyea DA, Kohrt HE, Medeiros BC (2011) Acute myeloid leukaemia in the elderly: a review. Br J Haematol 152:524–542. https://doi.org/10.1111/j.1365-2141.2010.08470.x

2. Rodrigues ACB da C, Costa RGA, Silva SLR, et al (2021) Cell signaling pathways as molecular targets to eliminate AML stem cells. Crit Rev Oncol Hematol 160:103277. https://doi.org/10.1016/j.critrevonc.2021.103277

3. Shallis RM, Wang R, Davidoff A, et al (2019) Epidemiology of acute myeloid leukemia: Recent progress and enduring challenges. Blood Rev 36:70–87. https://doi.org/10.1016/j.blre.2019.04.005

4. Lindsley RC, Mar BG, Mazzola E, et al (2015) Acute myeloid leukemia ontogeny is defined by distinct somatic mutations. Blood 125:1367–1376. https://doi.org/10.1182/blood-2014-11-610543

5. Bachas C, Schuurhuis GJ, Hollink IHIM, et al (2010) High-frequency type I/II mutational shifts between diagnosis and relapse are associated with outcome in pediatric AML: implications for personalized medicine. Blood 116:2752–2758. https://doi.org/10.1182/blood-2010-03-276519

6. Marjanovic I, Kostic J, Stanic B, et al (2016) Parallel targeted next generation sequencing of childhood and adult acute myeloid leukemia patients reveals uniform genomic profile of the disease. Tumour Biol J Int Soc Oncodevelopmental Biol Med 37:13391–13401. https://doi.org/10.1007/s13277-016-5142-7

7. Zjablovskaja P, Florian MC (2019) Acute Myeloid Leukemia: Aging and Epigenetics. Cancers 12:. https://doi.org/10.3390/cancers12010103

8. Arber DA, Orazi A, Hasserjian R, et al (2016) The 2016 revision to the World Health Organization classification of myeloid neoplasms and acute leukemia. Blood 127:2391–2405. https://doi.org/10.1182/blood-2016-03-643544

9. Grimwade D, Walker H, Oliver F, et al (1998) The importance of diagnostic cytogenetics on outcome in AML: analysis of 1,612 patients entered into the MRC AML 10 trial. The Medical Research Council Adult and Children’s Leukaemia Working Parties. Blood 92:2322–2333

10. Döhner H, Estey EH, Amadori S, et al (2010) Diagnosis and management of acute myeloid leukemia in adults: recommendations from an international expert panel, on behalf of the European LeukemiaNet. Blood 115:453–474. https://doi.org/10.1182/blood-2009-07-235358

11. Döhner H, Estey E, Grimwade D, et al (2017) Diagnosis and management of AML in adults: 2017 ELN recommendations from an international expert panel. Blood 129:424–447. https://doi.org/10.1182/blood-2016-08-733196

12. Mendez LM, Posey RR, Pandolfi PP (2019) The Interplay Between the Genetic and Immune Landscapes of AML: Mechanisms and Implications for Risk Stratification and Therapy. Front Oncol 9:1162. https://doi.org/10.3389/fonc.2019.01162

13. Sendker S, Reinhardt D, Niktoreh N (2021) Redirecting the Immune Microenvironment in Acute Myeloid Leukemia. Cancers 13:. https://doi.org/10.3390/cancers13061423

14. Basilico S, Wang X, Kennedy A, et al (2020) Dissecting the early steps of MLL induced leukaemogenic transformation using a mouse model of AML. Nat Commun 11:1407. https://doi.org/10.1038/s41467-020-15220-0

15. Levis M, Murphy KM, Pham R, et al (2005) Internal tandem duplications of the FLT3 gene are present in leukemia stem cells. Blood 106:673–680. https://doi.org/10.1182/blood-2004-05-1902

16. Karantanos T, Jones RJ (2019) Acute Myeloid Leukemia Stem Cell Heterogeneity and Its Clinical Relevance. Adv Exp Med Biol 1139:153–169. https://doi.org/10.1007/978-3-030-14366-4_9

17. Kokkaliaris KD, Scadden DT (2020) Cell interactions in the bone marrow microenvironment affecting myeloid malignancies. Blood Adv 4:3795–3803. https://doi.org/10.1182/bloodadvances.2020002127

18. Walkley CR, Olsen GH, Dworkin S, et al (2007) A microenvironment-induced myeloproliferative syndrome caused by retinoic acid receptor gamma deficiency. Cell 129:1097–1110. https://doi.org/10.1016/j.cell.2007.05.014

19. Lefebvre P, Thomas G, Gourmel B, et al (1991) Pharmacokinetics of oral all-trans retinoic acid in patients with acute promyelocytic leukemia. Leukemia 5:1054–1058

20. Johnson DE, Redner RL (2015) An ATRActive future for differentiation therapy in AML. Blood Rev 29:263–268. https://doi.org/10.1016/j.blre.2015.01.002

21. Bradbury DA, Aldington S, Zhu YM, Russell NH (1996) Down-regulation of bcl-2 in AML blasts by all-trans retinoic acid and its relationship to CD34 antigen expression. Br J Haematol 94:671–675. https://doi.org/10.1046/j.1365-2141.1996.d01-1838.x

22. Lehmann S, Bengtzen S, Broberg U, Paul C (2000) Effects of retinoids on cell toxicity and apoptosis in leukemic blast cells from patients with non-M3 AML. Leuk Res 24:19–25. https://doi.org/10.1016/s0145-2126(99)00153-8

23. Gudas LJ (2012) Emerging roles for retinoids in regeneration and differentiation in normal and disease states. Biochim Biophys Acta 1821:213–221. https://doi.org/10.1016/j.bbalip.2011.08.002

24. Kastan MB, Schlaffer E, Russo JE, et al (1990) Direct demonstration of elevated aldehyde dehydrogenase in human hematopoietic progenitor cells. Blood 75:1947–1950

25. Bidan N, Bailleul-Dubois J, Duval J, et al (2019) Transcriptomic Analysis of Breast Cancer Stem Cells and Development of a pALDH1A1:mNeptune Reporter System for Live Tracking. Proteomics 19:e1800454. https://doi.org/10.1002/pmic.201800454

26. Gasparetto M, Smith CA (2017) ALDHs in normal and malignant hematopoietic cells: Potential new avenues for treatment of AML and other blood cancers. Chem Biol Interact 276:46–51. https://doi.org/10.1016/j.cbi.2017.06.020

27. Schuurhuis GJ, Meel MH, Wouters F, et al (2013) Normal hematopoietic stem cells within the AML bone marrow have a distinct and higher ALDH activity level than co-existing leukemic stem cells. PloS One 8:e78897. https://doi.org/10.1371/journal.pone.0078897

28. Hoang VT, Buss EC, Wang W, et al (2015) The rarity of ALDH(+) cells is the key to separation of normal versus leukemia stem cells by ALDH activity in AML patients. Int J Cancer 137:525–536. https://doi.org/10.1002/ijc.29410

29. Ran D, Schubert M, Pietsch L, et al (2009) Aldehyde dehydrogenase activity among primary leukemia cells is associated with stem cell features and correlates with adverse clinical outcomes. Exp Hematol 37:1423–1434. https://doi.org/10.1016/j.exphem.2009.10.001

30. Blume R, Rempel E, Manta L, et al (2018) The molecular signature of AML with increased ALDH activity suggests a stem cell origin. Leuk Lymphoma 59:2201–2210. https://doi.org/10.1080/10428194.2017.1422862

31. Fan X, Molotkov A, Manabe S-I, et al (2003) Targeted disruption of Aldh1a1 (Raldh1) provides evidence for a complex mechanism of retinoic acid synthesis in the developing retina. Mol Cell Biol 23:4637–4648. https://doi.org/10.1128/mcb.23.13.4637-4648.2003

32. Xiao T, Shoeb M, Siddiqui MS, et al (2009) Molecular cloning and oxidative modification of human lens ALDH1A1: implication in impaired detoxification of lipid aldehydes. J Toxicol Environ Health A 72:577–584. https://doi.org/10.1080/15287390802706371

33. Wang B, Chen X, Wang Z, et al (2017) Aldehyde dehydrogenase 1A1 increases NADH levels and promotes tumor growth via glutathione/dihydrolipoic acid-dependent NAD+ reduction. Oncotarget 8:67043–67055. https://doi.org/10.18632/oncotarget.17688

34. Tomita H, Tanaka K, Tanaka T, Hara A (2016) Aldehyde dehydrogenase 1A1 in stem cells and cancer. Oncotarget 7:11018–11032. https://doi.org/10.18632/oncotarget.6920

35. Anderson DW, Schray RC, Duester G, Schneider JS (2011) Functional significance of aldehyde dehydrogenase ALDH1A1 to the nigrostriatal dopamine system. Brain Res 1408:81–87. https://doi.org/10.1016/j.brainres.2011.06.051

36. Fan H-H, Guo Q, Zheng J, et al (2021) ALDH1A1 Genetic Variations May Modulate Risk of Parkinson’s Disease in Han Chinese Population. Front Neurosci 15:620929. https://doi.org/10.3389/fnins.2021.620929

37. Choudhary S, Xiao T, Vergara LA, et al (2005) Role of aldehyde dehydrogenase isozymes in the defense of rat lens and human lens epithelial cells against oxidative stress. Invest Ophthalmol Vis Sci 46:259–267. https://doi.org/10.1167/iovs.04-0120

38. Lassen N, Bateman JB, Estey T, et al (2007) Multiple and additive functions of ALDH3A1 and ALDH1A1: cataract phenotype and ocular oxidative damage in Aldh3a1(-/-)/Aldh1a1(-/-) knock-out mice. J Biol Chem 282:25668–25676. https://doi.org/10.1074/jbc.M702076200

39. Levi BP, Yilmaz OH, Duester G, Morrison SJ (2009) Aldehyde dehydrogenase 1a1 is dispensable for stem cell function in the mouse hematopoietic and nervous systems. Blood 113:1670–1680. https://doi.org/10.1182/blood-2008-05-156752

40. Marjanovic I, Karan-Djurasevic T, Kostic T, et al (2020) Expression Pattern and Prognostic Significance of EVI1 Gene in Adult Acute Myeloid Leukemia Patients with Normal Karyotype. Indian J Hematol Blood Transfus Off J Indian Soc Hematol Blood Transfus 36:292–299. https://doi.org/10.1007/s12288-019-01227-1

41. Kustikova OS, Schwarzer A, Stahlhut M, et al (2013) Activation of Evi1 inhibits cell cycle progression and differentiation of hematopoietic progenitor cells. Leukemia 27:1127–1138. https://doi.org/10.1038/leu.2012.355

42. Wu X, Wang H, Deng J, et al (2019) Prognostic significance of the EVI1 gene expression in patients with acute myeloid leukemia: a meta-analysis. Ann Hematol 98:2485–2496. https://doi.org/10.1007/s00277-019-03774-z

43. Kallifatidis G, Labsch S, Rausch V, et al (2011) Sulforaphane increases drug-mediated cytotoxicity toward cancer stem-like cells of pancreas and prostate. Mol Ther J Am Soc Gene Ther 19:188–195. https://doi.org/10.1038/mt.2010.216

44. Liu L, Salnikov AV, Bauer N, et al (2014) Triptolide reverses hypoxia-induced epithelial-mesenchymal transition and stem-like features in pancreatic cancer by NF-κB downregulation. Int J Cancer 134:2489–2503. https://doi.org/10.1002/ijc.28583

45. Rausch V, Liu L, Kallifatidis G, et al (2010) Synergistic activity of sorafenib and sulforaphane abolishes pancreatic cancer stem cell characteristics. Cancer Res 70:5004–5013. https://doi.org/10.1158/0008-5472.CAN-10-0066

46. Lambrou GI, Hatziagapiou K, Vlahopoulos S (2020) Inflammation and tissue homeostasis: the NF-κB system in physiology and malignant progression. Mol Biol Rep. https://doi.org/10.1007/s11033-020-05410-w

47. Yang F, Xu Y, Liu C, et al (2018) NF-κB/miR-223-3p/ARID1A axis is involved in Helicobacter pylori CagA-induced gastric carcinogenesis and progression. Cell Death Dis 9:12. https://doi.org/10.1038/s41419-017-0020-9

48. Yoshino J, Akiyama Y, Shimada S, et al (2020) Loss of ARID1A induces a stemness gene ALDH1A1 expression with histone acetylation in the malignant subtype of cholangiocarcinoma. Carcinogenesis 41:734–742. https://doi.org/10.1093/carcin/bgz179

49. Kuo H-P, Wang Z, Lee D-F, et al (2013) Epigenetic roles of MLL oncoproteins are dependent on NF-κB. Cancer Cell 24:423–437. https://doi.org/10.1016/j.ccr.2013.08.019

50. Prajoko YW, Aryandono T (2019) The Effect of P-Glycoprotein (P-gp), Nuclear Factor-Kappa B (Nf-κb), and Aldehyde Dehydrogenase-1 (ALDH-1) Expression on Metastases, Recurrence and Survival in Advanced Breast Cancer Patients. Asian Pac J Cancer Prev APJCP 20:1511–1518. https://doi.org/10.31557/APJCP.2019.20.5.1511

51. Lück SC, Russ AC, Du J, et al (2010) KIT mutations confer a distinct gene expression signature in core binding factor leukaemia. Br J Haematol 148:925–937. https://doi.org/10.1111/j.1365-2141.2009.08035.x

52. Nasri F, Sadeghi F, Behranvand N, et al (2020) Oridonin Could Inhibit Inflammation and T-cell Immunoglobulin and Mucin-3/Galectin-9 (TIM-3/Gal-9) Autocrine Loop in the Acute Myeloid Leukemia Cell Line (U937) as Compared to Doxorubicin. Iran J Allergy Asthma Immunol 19:602–611. https://doi.org/10.18502/ijaai.v19i6.4929

53. Burchert A (2021) Maintenance therapy for FLT3-ITD-mutated acute myeloid leukemia. Haematologica 106:664–670. https://doi.org/10.3324/haematol.2019.240747

54. Man CH, Fung TK, Ho C, et al (2012) Sorafenib treatment of FLT3-ITD(+) acute myeloid leukemia: favorable initial outcome and mechanisms of subsequent nonresponsiveness associated with the emergence of a D835 mutation. Blood 119:5133–5143. https://doi.org/10.1182/blood-2011-06-363960

55. Elcheva IA, Wood T, Chiarolanzio K, et al (2020) RNA-binding protein IGF2BP1 maintains leukemia stem cell properties by regulating HOXB4, MYB, and ALDH1A1. Leukemia 34:1354–1363. https://doi.org/10.1038/s41375-019-0656-9

56. Allison SE, Chen Y, Petrovic N, et al (2017) Activation of ALDH1A1 in MDA-MB-468 breast cancer cells that over-express CYP2J2 protects against paclitaxel-dependent cell death mediated by reactive oxygen species. Biochem Pharmacol 143:79–89. https://doi.org/10.1016/j.bcp.2017.07.020

57. Bogen A, Buske C, Hiddemann W, et al (2017) Variable aldehyde dehydrogenase activity and effects on chemosensitivity of primitive human leukemic cells. Exp Hematol 47:54–63. https://doi.org/10.1016/j.exphem.2016.10.012

58. Venton G, Pérez-Alea M, Baier C, et al (2016) Aldehyde dehydrogenases inhibition eradicates leukemia stem cells while sparing normal progenitors. Blood Cancer J 6:e469. https://doi.org/10.1038/bcj.2016.78

59. Gasparetto M, Pei S, Minhajuddin M, et al (2017) Targeted therapy for a subset of acute myeloid leukemias that lack expression of aldehyde dehydrogenase 1A1. Haematologica 102:1054–1065. https://doi.org/10.3324/haematol.2016.159053

60. Barrett T, Wilhite SE, Ledoux P, et al (2013) NCBI GEO: archive for functional genomics data sets--update. Nucleic Acids Res 41:D991–995. https://doi.org/10.1093/nar/gks1193

61. Deng M, Brägelmann J, Kryukov I, et al (2017) FirebrowseR: an R client to the Broad Institute’s Firehose Pipeline. Database J Biol Databases Curation 2017:. https://doi.org/10.1093/database/baw160

62. Jensen MA, Ferretti V, Grossman RL, Staudt LM (2017) The NCI Genomic Data Commons as an engine for precision medicine. Blood 130:453–459. https://doi.org/10.1182/blood-2017-03-735654

63. Gao J, Aksoy BA, Dogrusoz U, et al (2013) Integrative analysis of complex cancer genomics and clinical profiles using the cBioPortal. Sci Signal 6:pl1. https://doi.org/10.1126/scisignal.2004088

64. Robinson MD, Oshlack A (2010) A scaling normalization method for differential expression analysis of RNA-seq data. Genome Biol 11:R25. https://doi.org/10.1186/gb-2010-11-3-r25

65. Ritchie ME, Phipson B, Wu D, et al (2015) limma powers differential expression analyses for RNA-sequencing and microarray studies. Nucleic Acids Res 43:e47. https://doi.org/10.1093/nar/gkv007

66. Rice KL, Izon DJ, Ford J, et al (2008) Overexpression of stem cell associated ALDH1A1, a target of the leukemogenic transcription factor TLX1/HOX11, inhibits lymphopoiesis and promotes myelopoiesis in murine hematopoietic progenitors. Leuk Res 32:873–883. https://doi.org/10.1016/j.leukres.2007.11.001

67. Li Z, Huang H, Li Y, et al (2012) Up-regulation of a HOXA-PBX3 homeobox-gene signature following down-regulation of miR-181 is associated with adverse prognosis in patients with cytogenetically abnormal AML. Blood 119:2314–2324. https://doi.org/10.1182/blood-2011-10-386235

68. Ignatz-Hoover JJ, Wang V, Mackowski NM, et al (2018) Aberrant GSK3β nuclear localization promotes AML growth and drug resistance. Blood Adv 2:2890–2903. https://doi.org/10.1182/bloodadvances.2018016006

69. Reinisch A, Chan SM, Thomas D, Majeti R (2015) Biology and Clinical Relevance of Acute Myeloid Leukemia Stem Cells. Semin Hematol 52:150–164. https://doi.org/10.1053/j.seminhematol.2015.03.008

70. Wang Z, Chen J, Wang M, et al (2021) One Stone, Two Birds: The Roles of Tim-3 in Acute Myeloid Leukemia. Front Immunol 12:618710. https://doi.org/10.3389/fimmu.2021.618710

71. Kikushige Y, Miyamoto T, Yuda J, et al (2015) A TIM-3/Gal-9 Autocrine Stimulatory Loop Drives Self-Renewal of Human Myeloid Leukemia Stem Cells and Leukemic Progression. Cell Stem Cell 17:341–352. https://doi.org/10.1016/j.stem.2015.07.011

72. Birkenkamp KU, Geugien M, Schepers H, et al (2004) Constitutive NF-kappaB DNA-binding activity in AML is frequently mediated by a Ras/PI3-K/PKB-dependent pathway. Leukemia 18:103–112. https://doi.org/10.1038/sj.leu.2403145

73. Takahashi S, Harigae H, Ishii KK, et al (2005) Over-expression of Flt3 induces NF-kappaB pathway and increases the expression of IL-6. Leuk Res 29:893–899. https://doi.org/10.1016/j.leukres.2005.01.008

74. Imbert V, Peyron J-F (2017) NF-κB in Hematological Malignancies. Biomedicines 5:. https://doi.org/10.3390/biomedicines5020027

75. Grosjean-Raillard J, Adès L, Boehrer S, et al (2008) Flt3 receptor inhibition reduces constitutive NFkappaB activation in high-risk myelodysplastic syndrome and acute myeloid leukemia. Apoptosis Int J Program Cell Death 13:1148–1161. https://doi.org/10.1007/s10495-008-0243-4

76. Zhou J, Ching YQ, Chng W-J (2015) Aberrant nuclear factor-kappa B activity in acute myeloid leukemia: from molecular pathogenesis to therapeutic target. Oncotarget 6:5490–5500. https://doi.org/10.18632/oncotarget.3545

77. Varisli L, Cen O, Vlahopoulos S (2019) Dissecting pharmacological effects of Chloroquine in cancer treatment: interference with inflammatory signaling pathways. Immunology. https://doi.org/10.1111/imm.13160

78. Kulkarni U, Mathews V (2021) Evolving Chemotherapy Free Regimens for Acute Promyelocytic Leukemia. Front Oncol 11:621566. https://doi.org/10.3389/fonc.2021.621566

79. Kulkarni U, Ganesan S, Alex AA, et al (2020) A phase II study evaluating the role of bortezomib in the management of relapsed acute promyelocytic leukemia treated upfront with arsenic trioxide. Cancer Med 9:2603–2610. https://doi.org/10.1002/cam4.2883

80. Takahashi S (2020) Current Understandings of Myeloid Differentiation Inducers in Leukemia Therapy. Acta Haematol 1–9. https://doi.org/10.1159/000510980

81. Sreenivasan Y, Sarkar A, Manna SK (2003) Mechanism of cytosine arabinoside-mediated apoptosis: role of Rel A (p65) dephosphorylation. Oncogene 22:4356–4369. https://doi.org/10.1038/sj.onc.1206486

82. Murphy T, Yee KWL (2017) Cytarabine and daunorubicin for the treatment of acute myeloid leukemia. Expert Opin Pharmacother 18:1765–1780. https://doi.org/10.1080/14656566.2017.1391216

83. Löwenberg B (2013) Sense and nonsense of high-dose cytarabine for acute myeloid leukemia. Blood 121:26–28. https://doi.org/10.1182/blood-2012-07-444851

84. Zhong Y, Qiu R-Z, Sun S-L, et al (2020) Small-Molecule Fms-like Tyrosine Kinase 3 Inhibitors: An Attractive and Efficient Method for the Treatment of Acute Myeloid Leukemia. J Med Chem 63:12403–12428. https://doi.org/10.1021/acs.jmedchem.0c00696

85. Mosquera Orgueira A, Bao Pérez L, Mosquera Torre A, et al (2020) FLT3 inhibitors in the treatment of acute myeloid leukemia: current status and future perspectives. Minerva Med 111:427–442. https://doi.org/10.23736/S0026-4806.20.06989-X

86. Griessinger E, Frelin C, Cuburu N, et al (2008) Preclinical targeting of NF-kappaB and FLT3 pathways in AML cells. Leukemia 22:1466–1469. https://doi.org/10.1038/sj.leu.2405102

87. Wang C, Lu J, Wang Y, et al (2012) Combined effects of FLT3 and NF-κB selective inhibitors on acute myeloid leukemia in vivo. J Biochem Mol Toxicol 26:35–43. https://doi.org/10.1002/jbt.20411

88. Vlahopoulos SA (2017) Aberrant control of NF-κB in cancer permits transcriptional and phenotypic plasticity, to curtail dependence on host tissue: molecular mode. Cancer Biol Med 14:254–270. https://doi.org/10.20892/j.issn.2095-3941.2017.0029

89. Yang W, Xie J, Hou R, et al (2020) Disulfiram/cytarabine eradicates a subset of acute myeloid leukemia stem cells with high aldehyde dehydrogenase expression. Leuk Res 92:106351. https://doi.org/10.1016/j.leukres.2020.106351

90. Jacquel A, Luciano F, Robert G, Auberger P (2018) Implication and Regulation of AMPK during Physiological and Pathological Myeloid Differentiation. Int J Mol Sci 19:. https://doi.org/10.3390/ijms19102991

91. Strobl H, Knapp W (2004) Myeloid cell-associated lysosomal proteins as flow cytometry markers for leukocyte lineage classification. J Biol Regul Homeost Agents 18:335–339

92. Jang JE, Eom J-I, Jeung H-K, et al (2017) AMPK-ULK1-Mediated Autophagy Confers Resistance to BET Inhibitor JQ1 in Acute Myeloid Leukemia Stem Cells. Clin Cancer Res Off J Am Assoc Cancer Res 23:2781–2794. https://doi.org/10.1158/1078-0432.CCR-16-1903

93. Rothe K, Porter V, Jiang X (2019) Current Outlook on Autophagy in Human Leukemia: Foe in Cancer Stem Cells and Drug Resistance, Friend in New Therapeutic Interventions. Int J Mol Sci 20:. https://doi.org/10.3390/ijms20030461

94. Bosc C, Broin N, Fanjul M, et al (2020) Autophagy regulates fatty acid availability for oxidative phosphorylation through mitochondria-endoplasmic reticulum contact sites. Nat Commun 11:4056. https://doi.org/10.1038/s41467-020-17882-2

95. Wu Z, Shen L, Inatomi Y, et al (2003) Effects of TNFalpha on the growth and sensitivity to cytosine arabinoside of blast progenitors in acute myelogenous leukemia with special reference to the role of NF-kappaB. Leuk Res 27:1009–1018. https://doi.org/10.1016/s0145-2126(03)00069-9

96. Altman JK, Szilard A, Goussetis DJ, et al (2014) Autophagy is a survival mechanism of acute myelogenous leukemia precursors during dual mTORC2/mTORC1 targeting. Clin Cancer Res Off J Am Assoc Cancer Res 20:2400–2409. https://doi.org/10.1158/1078-0432.CCR-13-3218

97. Piao S, Ojha R, Rebecca VW, et al (2017) ALDH1A1 and HLTF modulate the activity of lysosomal autophagy inhibitors in cancer cells. Autophagy 13:2056–2071. https://doi.org/10.1080/15548627.2017.1377377

98. Visser N, Lourens HJ, Huls G, et al (2021) Inhibition of Autophagy Does Not Re-Sensitize Acute Myeloid Leukemia Cells Resistant to Cytarabine. Int J Mol Sci 22:. https://doi.org/10.3390/ijms22052337

99. Zhou F-L, Zhang W-G, Wei Y-C, et al (2010) Involvement of oxidative stress in the relapse of acute myeloid leukemia. J Biol Chem 285:15010–15015. https://doi.org/10.1074/jbc.M110.103713

100. Sallmyr A, Fan J, Datta K, et al (2008) Internal tandem duplication of FLT3 (FLT3/ITD) induces increased ROS production, DNA damage, and misrepair: implications for poor prognosis in AML. Blood 111:3173–3182. https://doi.org/10.1182/blood-2007-05-092510

101. Rebechi MT, Pratz KW (2017) Genomic instability is a principle pathologic feature of FLT3 ITD kinase activity in acute myeloid leukemia leading to clonal evolution and disease progression. Leuk Lymphoma 58:1–11. https://doi.org/10.1080/10428194.2017.1283031

102. Robinson AJ, Davies S, Darley RL, Tonks A (2021) Reactive Oxygen Species Rewires Metabolic Activity in Acute Myeloid Leukemia. Front Oncol 11:632623. https://doi.org/10.3389/fonc.2021.632623

103. Hao W, Qi T, Pan L, et al (2018) Effects of the stimuli-dependent enrichment of 8-oxoguanine DNA glycosylase1 on chromatinized DNA. Redox Biol 18:43–53. https://doi.org/10.1016/j.redox.2018.06.002

104. Vlahopoulos S, Adamaki M, Khoury N, et al (2018) Roles of DNA repair enzyme OGG1 in innate immunity and its significance for lung cancer. Pharmacol Ther. https://doi.org/10.1016/j.pharmthera.2018.09.004

105. Moto M, Okamura M, Muto T, et al (2005) Molecular pathological analysis on the mechanism of liver carcinogenesis in dicyclanil-treated mice. Toxicology 207:419–436. https://doi.org/10.1016/j.tox.2004.10.011

106. Liddiard K, Hills R, Burnett AK, et al (2010) OGG1 is a novel prognostic indicator in acute myeloid leukaemia. Oncogene 29:2005–2012. https://doi.org/10.1038/onc.2009.462

107. Owen N, Minko IG, Moellmer SA, et al (2021) Enhanced cytarabine-induced killing in OGG1-deficient acute myeloid leukemia cells. Proc Natl Acad Sci U S A 118:. https://doi.org/10.1073/pnas.2016833118

108. Zhou AL, Wang X, Yu W, et al (2020) Expression level of PD-L1 is involved in ALDH1A1-mediated poor prognosis in patients with head and neck squamous cell carcinoma. Pathol Res Pract 216:153093. https://doi.org/10.1016/j.prp.2020.153093

109. Castagnoli L, Cancila V, Cordoba-Romero SL, et al (2019) WNT signaling modulates PD-L1 expression in the stem cell compartment of triple-negative breast cancer. Oncogene 38:4047–4060. https://doi.org/10.1038/s41388-019-0700-2

110. Hwang H-S, Han A-R, Lee JY, et al (2019) Enhanced Anti-Leukemic Effects through Induction of Immunomodulating Microenvironment by Blocking CXCR4 and PD-L1 in an AML Mouse Model. Immunol Invest 48:96–105. https://doi.org/10.1080/08820139.2018.1497057

111. Dong Y, Han Y, Huang Y, et al (2020) PD-L1 Is Expressed and Promotes the Expansion of Regulatory T Cells in Acute Myeloid Leukemia. Front Immunol 11:1710. https://doi.org/10.3389/fimmu.2020.01710

112. Taghiloo S, Asgarian-Omran H (2021) Immune evasion mechanisms in acute myeloid leukemia: A focus on immune checkpoint pathways. Crit Rev Oncol Hematol 157:103164. https://doi.org/10.1016/j.critrevonc.2020.103164

113. Yang S-M, Martinez NJ, Yasgar A, et al (2018) Discovery of Orally Bioavailable, Quinoline-Based Aldehyde Dehydrogenase 1A1 (ALDH1A1) Inhibitors with Potent Cellular Activity. J Med Chem 61:4883–4903. https://doi.org/10.1021/acs.jmedchem.8b00270

114. Yasgar A, Titus SA, Wang Y, et al (2017) A High-Content Assay Enables the Automated Screening and Identification of Small Molecules with Specific ALDH1A1-Inhibitory Activity. PloS One 12:e0170937. https://doi.org/10.1371/journal.pone.0170937

115. Ma Z, Jiang L, Li G, et al (2020) Design, synthesis of 1,3-dimethylpyrimidine-2,4-diones as potent and selective aldehyde dehydrogenase 1A1 (ALDH1A1) inhibitors with glucose consumption improving activity. Bioorganic Chem 101:103971. https://doi.org/10.1016/j.bioorg.2020.103971

116. Li B, Yang K, Liang D, et al (2021) Discovery and development of selective aldehyde dehydrogenase 1A1 (ALDH1A1) inhibitors. Eur J Med Chem 209:112940. https://doi.org/10.1016/j.ejmech.2020.112940

