## Supplement for "Lower RNA expression of ALDH1A1 distinguishes the favorable risk group in acute myeloid leukemia"

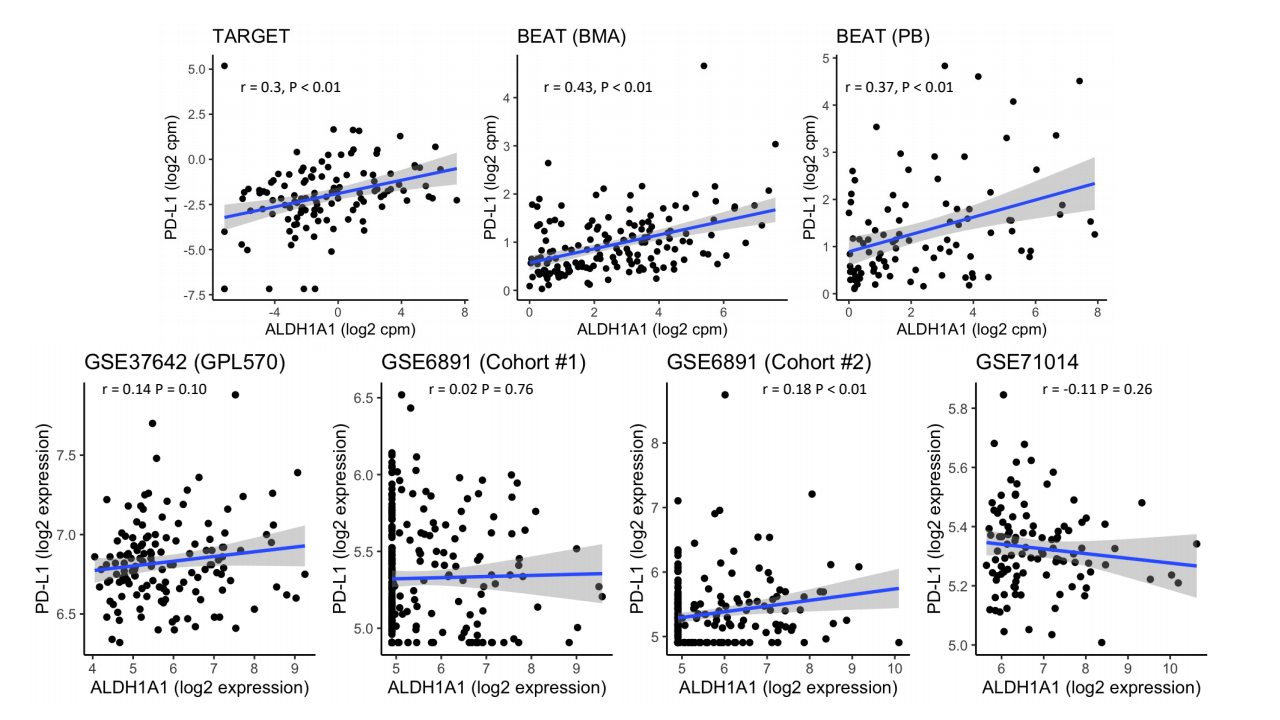

**Figure S1. Correlation between ALDH1A1 and PD-L1 expression in patient cohorts.** Scatterplots of expression values are generated for TARGET (N = 119), BEAT (BMA) (N = 163), BEAT (PB) (N = 95), GSE37642 (GPL570) (N = 140), GSE6891 (Cohort #1) (N = 247) and GSE6891 (Cohort#2) (N = 214) cohorts. Additional cohorts were not included because of low gene counts (TCGA) or because PD-L1 was not profiled (GSE37642 (GPL96)). For each cohort, the Pearson correlation (*r*) and p-value are reported. Each blue line is the linear regression line for each scatterplot with the gray region denoting the 95% confidence interval.
